# Electronic early notification of sepsis in hospitalized ward patients: a study protocol for a stepped-wedge cluster randomized controlled trial

**DOI:** 10.1101/2021.05.20.21257511

**Authors:** Yaseen M Arabi, Abdulmohsen Al Saawi, Mohammed Al Zahrani, Ali Al Khathaami, Raed H AlHazme, Abdullah Al Mutrafy, Ali Al Qarni, Ahmed Al Shouabi, Eman Al Qasim, Sheryl Ann Abdukahil, Fawaz Q Al-Rabeeah, Huda Al Ghamdi, Ebtisam Al Ghamdi, Mariam Alansari, Khadega A Abuelgasim, Abdulaleem Alattasi, John Alchin, Hasan M Al-Dorzi, Abdulaziz A Ghamdi, Fahad Al-Hameed, Ahmad Alharbi, Mohamed Hussein, Wasil Jastaniah, Mufareh Edah AlKatheri, Hassan AlMarhabi, Hani T Mustafa, Joan Jones, Saad Al-Qahtani, Shaher Qahtani, Ahmad S Qureshi, Salih Bin Salih, Nahar Alselaim, Nabeeha Tashkandi, Ramesh Kumar Vishwakarma, Emad AlWafi, Ali H Alyami, Zeyad Al Yousef, for the SCREEN Trial Group

## Abstract

**Background:** To examine the effect of screening for sepsis using an electronic sepsis alert versus no alert in hospitalized patients admitted to wards on hospital mortality.

**Methods:** This study is conducted in 45 medical-surgical-oncology wards in five hospitals. Based on the quick Sequential Organ Failure Assessment (qSOFA), an electronic alert has been developed in the hospital Electronic Medical Record system. The alert system sends notifications of “Possible Sepsis Alert” to the bedside nurse, charge nurse, and primary medical team and requires an acknowledgment in the health information system from the bedside nurse and physician. In addition, data on the alert are displayed on management dashboards for each ward. Initially, all wards had a masked alert for 2 months. Hospital wards are then allocated in a randomized fashion to either active or masked alert, such that the alert is activated in five new randomly selected wards every two months until all wards have the active alert. The primary endpoint is in-hospital mortality by 90 days.

**Discussion:** The trial has started in October 2019 and is expected to continue for 22 months enrolling more than 62550 hospitalized patients.

**Trial registration:** ClinicalTrials.gov NCT04078594. Registered on September 6, 2019, https://clinicaltrials.gov/ct2/show/NCT04078594

## Background

Sepsis is a major cause of morbidity and mortality among hospitalized patients and the outcome of affected patients is greatly dependent on the time-sensitive administration of appropriate antimicrobials, fluid resuscitation, and source control.(1, 2) Using machine learning analytics, a retrospective multicenter study evaluated the impact of delaying the implementation of the Surviving Sepsis Campaign 3-hour bundle elements in 5,072 adult patients with severe sepsis or septic shock.(3) The study found that the time delays significantly associated with mortality were 20 minutes for serum lactate, 50 minutes for blood culture, 100 minutes for intravenous crystalloids, and 125 minutes for antibiotic therapy.(3) Unfortunately, studies have shown that these elements of sepsis management are often delayed because of delayed sepsis recognition.(4-6)

To facilitate early sepsis recognition, the Surviving Sepsis Campaign guidelines have recommended that hospitals have a performance improvement program for sepsis, including sepsis screening for acutely ill, high-risk patients.(7) Several observational studies have demonstrated that screening patients on hospital wards for sepsis was associated with improved processes of care and reduced mortality. A study from our center found that the implementation of a multifaceted intervention including sepsis electronic⍰alert (e-alert) with sepsis response team was associated with earlier identification of sepsis, increase in compliance with sepsis resuscitation bundle, and reduction in the need for mechanical ventilation and in-hospital mortality and length of stay.(8) A pre and post study evaluated a system using a triad of real-time electronic surveillance, mobile alerts received by nurses for all positive sepsis screenings as well as severe sepsis and shock alerts, and specific decision support for early goal-directed therapy.(9) During the study period, sepsis-related mortality decreased by 53% from 90 to 42 deaths per 1000 sepsis cases (p=0.03) and the 30-day readmission rate by 30.8% from 19.1% to 13.2% (p=0.05).(9) A prospective observational study compared a real-time sepsis alert with no alert in 6 medical wards in a 1250-bed academic medical center.(10) The study found that within 12 hours of the sepsis alert, 70.8% of patients in the intervention group had received ≥1 intervention vs. 55.8% in the control group (p=0.02).(10) Antibiotic escalation, intravenous fluid administration, oxygen therapy, and diagnostic tests were all increased in the intervention group.(10) More recently, a randomized controlled trial (RCT) at two medical-surgical intensive care units (ICUs) compared an electronic sepsis alert with the same alert combined with a machine learning algorithm (experimental group).(11) On receiving an alert, the care team evaluated the patient and initiated the severe sepsis bundle, if appropriate.(11) The length of stay decreased from 13.0 days in the control group to 10.3 days in the experimental group (p=0.04) and hospital mortality by 12.4% (p=0.02).(11) A before-after study evaluated the impact of a multidisciplinary sepsis education program targeting sepsis recognition by using both the quick Sequential Organ Failure Assessment (qSOFA) and organ dysfunction criteria with nurse empowerment to activate the rapid response team (RRT) when sepsis was suspected.(12) The study found that the time to recognition (qSOFA-to-RRT) improved from a median of 11.8 hours (interquartile range: 3.4, 34.3) pre to 1.7 hours (interquartile range: 0, 11.7) post (p=0.005).(12) Time from qSOFA to antibiotics improved from 1.4 hours (interquartile range:2.4, 6.2) pre to -4.7 hours (interquartile range:-25.4, 1.8) post (p <0.01).(12) Using qSOFA, compliance improved for antibiotics administration within 3 hours from 60% pre to 87% post (p=0.02).(12) A systematic review of 16 observational studies found that digital alert systems for sepsis were associated with a reduction in length of hospital stay.(13)

On the other hand, some data questioned the benefit of sepsis screening among hospitalized patients. A systematic review of six studies of electronic and paper-based sepsis screening suggested improvement in care processes on the use of diagnostics.(14) However, the effect on treatment (fluid resuscitation and antibiotic administration) and outcome measures was less consistent.(14) Most existing studies are pre-post intervention studies,(15-24) and showed a large reduction in mortality even with modest improvement in the compliance with the bundle. One of the potential explanations is that early detection leads to the inclusion of mild cases (ascertainment bias) and therefore artificially improving mortality rates. Therefore, the observed effect may be exaggerated or even maybe solely related to this phenomenon. Two RCTs showed no benefit of screening for sepsis in the ICU setting. A trial in a medical ICU (n= 442) showed that electronic screening using modified systemic inflammatory response syndrome versus usual care was not associated with a difference in time to new antibiotics, amount of fluid administered, ICU length of stay, hospital length of stay, and mortality.(25) Another trial in medical-surgical ICU patients (n=417) found no difference between the electronic tool and usual care in the primary outcome of time to completion of all indicated Surviving Sepsis Campaign 6-hour Sepsis Resuscitation Bundle elements, ICU mortality, ICU-free days, and ventilator-free days.(26) Besides, RCTs that tested real-time electronic alerts for clinical deterioration, which included sepsis in ward patients, found no significant difference in outcomes.(27, 28) Finally, screening for sepsis may lead to unintended consequences, such as overtreatment. One study found that the implementation of an electronic sepsis alert was associated with increases in antibiotic use and health care facility–onset Clostridium difficile infection.(29)

Therefore, it is unclear whether screening for sepsis using an electronic alert in hospitalized patients improves outcomes. The true effect of these interventions remains uncertain and an RCT is needed. The objective of the Stepped-wedge Cluster Randomized Controlled Trial of Electronic Early Notification of Sepsis in Hospitalized Ward Patients (SCREEN Trial) is to examine the effect of electronic screening for sepsis compared to no screening among hospitalized ward patients on all-cause 90-day hospital mortality.

## Methods

### Settings

The study is conducted in the 5 Ministry of National Guard Health Affairs (MNGHA) hospitals (King Abdulaziz Medical City-Riyadh, King Abdulaziz Medical City-Jeddah, and Prince Mohammed Bin Abdul Aziz Hospital – Al Madinah, King Abdulaziz Hospital - Al Ahsa, and Imam Abdulrahman Al Faisal Hospital – Dammam) (**Supplementary file Table S1**). All MNGHA hospitals have an integrated Electronic Medical Record (EMR) system, BESTCare, which has been implemented as a joint venture with the Seoul National University Bundang Hospital, South Korea. Its critical applications include the clinical documentation, computerized physician order entry, clinical decision support system, and clinical data warehouse. The system is also interfaced with the hospital’s vital signs measurement devices.

### Study Design

The SCREEN trial is designed as a stepped-wedge cluster RCT. Hospital wards are randomized to have active sepsis alert versus masked alert in phases. The study consists of nine periods of two-month steps with a phased introduction of the intervention. Initially, all wards had a masked alert for 2 months (see the section on sample size calculation). At each step, the alert is activated in five new randomly selected wards, until all wards eventually have the active alert (**Figure 1**). This design is suitable for quality improvement projects and allows comparison with the control group as well as over time. In addition, by having the masked alert, an additional comparison between patients with masked and active alerts will be performed. A SPIRIT figure is included as **Figure 2**, with a checklist included as a supplementary file.

**Figure 1:**
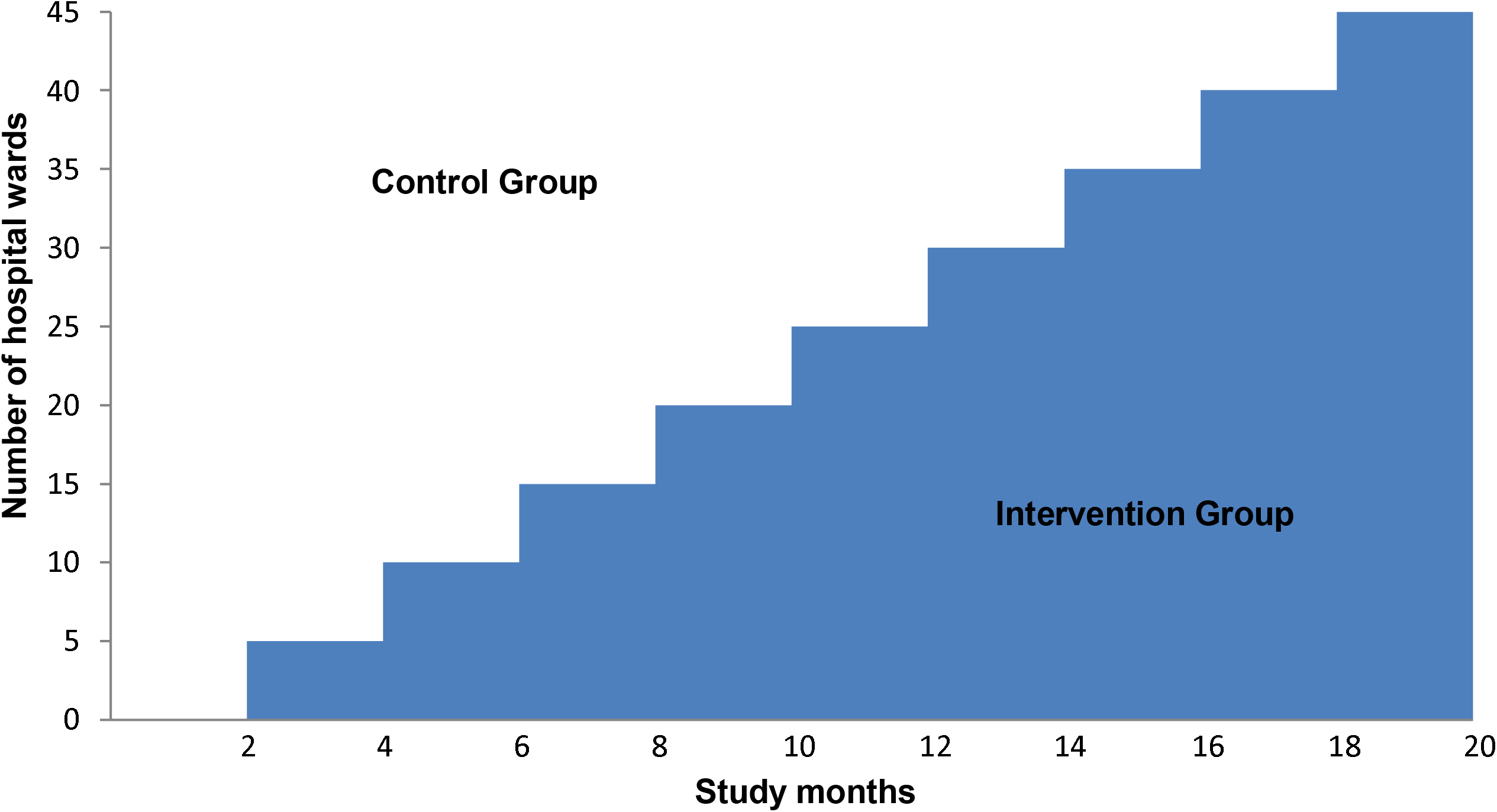
SCREEN trial design as a stepped-wedge cluster randomized trial.

**Figure 2:**
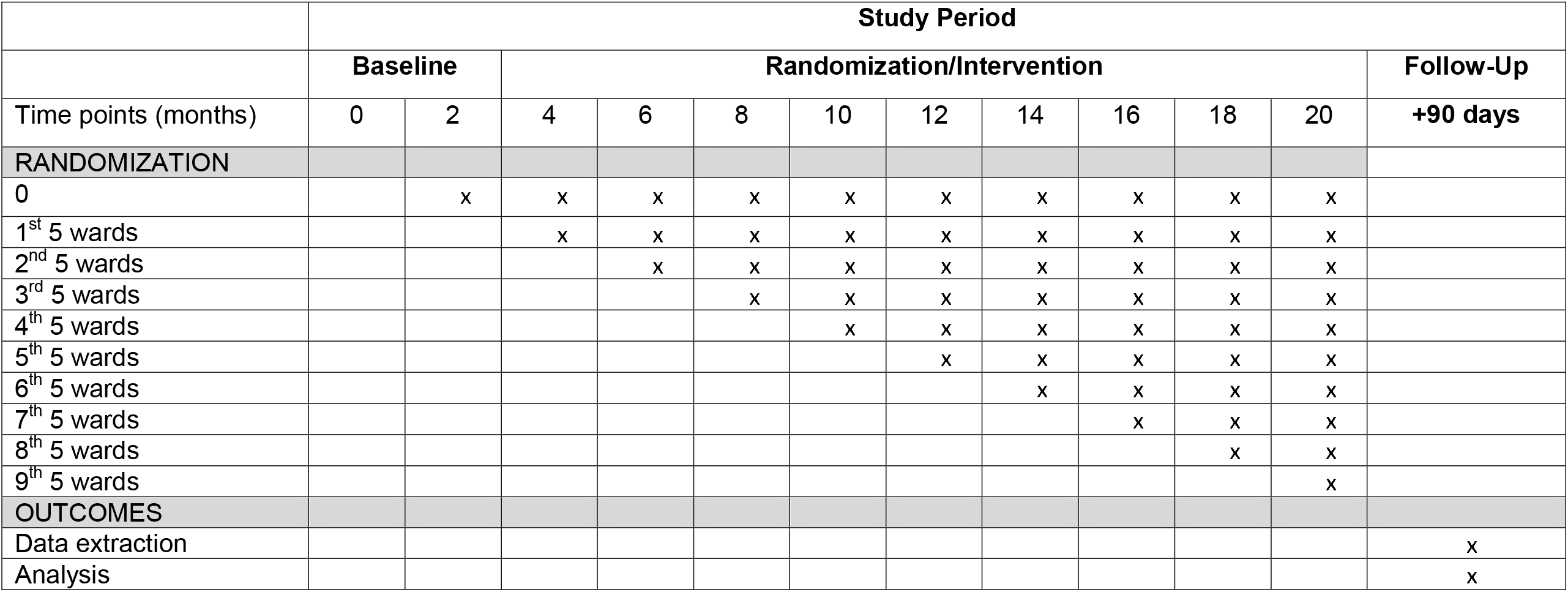
SPIRIT Figure for the SCREEN trial. Standard Protocol Items: Recommendations for Interventional Trials (SPIRIT) schedule.

This study was approved by the MNGHA Institutional Review Board (IRB). The study has been registered in clinicaltrials.gov (NCT0407859).

### Inclusion and Exclusion Criteria

#### Ward level inclusion and exclusion criteria

##### Inclusion Criteria

Inpatient wards, defined as wards used to manage inpatients, in the five MNGHA hospitals.

##### Exclusion Criteria

1. Cardiology, transplant, pediatric, obstetric wards
2. ICUs and emergency departments
3. Operating rooms
4. Outpatient clinics
5. Daycare wards, endoscopy, outpatient procedure areas, hemodialysis units

#### Patient-level inclusion and exclusion criteria

##### Inclusion Criteria

1. Aged 14 years or older
2. Checked in as inpatient status to one of the study wards

##### Exclusion Criteria

No commitment for full life support at the time of arrival to the study ward (designated as Do-Not-Resuscitate status)

### Interventions

#### The sepsis alert

In the study wards, vital signs are logged into the electronic medical record (BESTCare system) every 4 hours, and the Glasgow Coma Scale (GCS) every 12 hours. We developed a sepsis alert in the hospital information system based on qSOFA. The alert works as follows (**Figure 3**):

**Figure 3:**
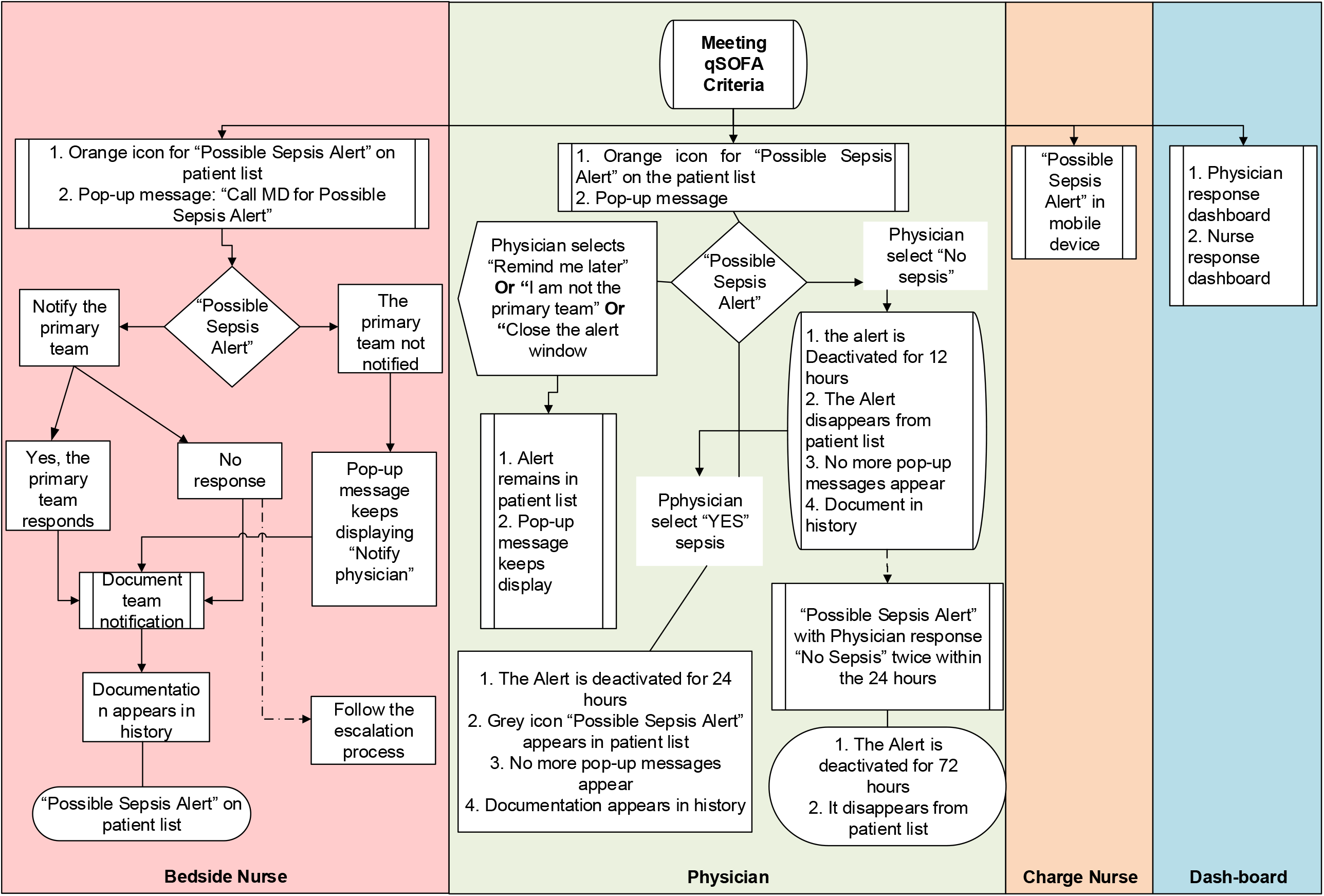
Workflow of the electronic alert.

1. The screening tool automatically scans the documented vital signs and GCS and assigns one point for low blood pressure (systolic blood pressure ≤ 100 mmHg), increased respiratory rate (≥ 22 breaths/ min), or altered mentation (Glasgow Coma Scale < 15).
2. If the total score is 2 or more based on the above vital signs documented within 12 hours, the system generates an electronic alert.
3. This alert results in the following:
  a. In the bedside nurse workflow: The alert appears in the nurse work list as “Possible Sepsis Alert” and a pop-up message appears to notify the primary medical team and to document that a physician has received the message.
  b. In the treating physician workflow: A pop-up message appears on the physician screen when the patient’s file is accessed as “Possible Sepsis Alert”. The physician is asked to assess the patient’s medical record and document if the patient has sepsis or not. Other options include “remind me later” and “I am not from the primary team”. By responding “Yes”, the sepsis alert is closed and the system is deactivated for 24 hours. By responding “No”, the system is deactivated for 12 hours only. By responding “remind me later” or “I am not from the primary team”, the pop-up message appears on each subsequent physician access of the file (**Figure 2**).
  c. The ward charge nurse gets a notification on a mobile device (iPod) that has a custom built application for an immediate alert with visual and sound notification.
  d. If the goals of care for a given patient are modified to Do-Not-Resuscitate, the system is deactivated for that patient.
4. Data are also be used to create a dashboard for each ward with the following indicators
  a. Number of alerts
  b. Percent of alerts with documented notification by nurse to the primary medical team and time to notification.
  c. Percent of alerts with documented assessment by physician and time to assessment.

Access to the dashboard is provided to all nursing and physician managers. Reports and feedback is provided every two weeks, with frequent meetings to review results and discuss approaches for improvement in the involved wards.

### Intervention Group (active alert)

In this group of wards, the alert is active, which means that the alert is visible to the treating team as described above. The alert prompts a nurse-to-physician communication and a physician assessment of patients.

### Control Group (masked alert)

In this group of wards, the alert will be inactive in EMR frontend for the treating team and will remain active in the backend of the EMR. Sepsis recognition is according to existing practice. This enables collecting data on the sepsis alert patients whether they are on active or masked control wards.

### Co-interventions

Hospital-wide sepsis awareness campaign in all 5 hospitals was started at the beginning of the study with training sessions open to all staff on the importance of timely interventions for sepsis. Project in-service training sessions are provided to the involved medical and nursing departments at the beginning of the project. Monthly webinars are conducted with active ward leaders. An intranet page was developed with educational resources (videos, presentations, documents, posters and related links) that explained this project and provided guidance. The medical management and assessment are at the discretion of the treating team. However, this project adopts the 2016 Surviving Sepsis Campaign guidelines and the hour-1 bundle, but without any specific monitoring of bundle compliance.(30, 31)

### Randomization

The five hospitals have collectively 46 eligible wards; two wards were merged due to similarities to create 9 clusters of 5 wards each. Wards are randomized into a total of 10 steps, one baseline and 9 implementation steps (**Figure 1**). The randomization list is maintained concealed by the study biostatistician from the study team and hospital teams. One month prior to each 5-ward implementation step, the allocation of the upcoming 5 wards is made open, so the ward staff prepare for the implementation.

### Study Cohorts

#### Intention-to-treat (ITT) cohort

This cohort includes all patients checked in to the candidate wards. This population constitutes the patients who are subjected to sepsis screening by the sepsis alert system, whether they develop an alert or not. The ITT analysis also implies that patients in the ITT cohort in the wards belonging to a particular randomization period will be analyzed in accordance with their planned randomization regardless of what happen during the trial. For example, if a ward was planned to receive an alert during a period and for technical reasons that alert system was not operational, patients recruited in that ward during that period will be analyzed as receiving the alert. The primary analysis will be based on this population.

#### Alert cohort

This cohort represents the subset of patients who had the alert.

## Data

### Data extraction

All data will be extracted from the EMR. Data will be extracted as per the definitions in **Tables 1, Supplementary file Table S4-S10**.

**Table 1:**
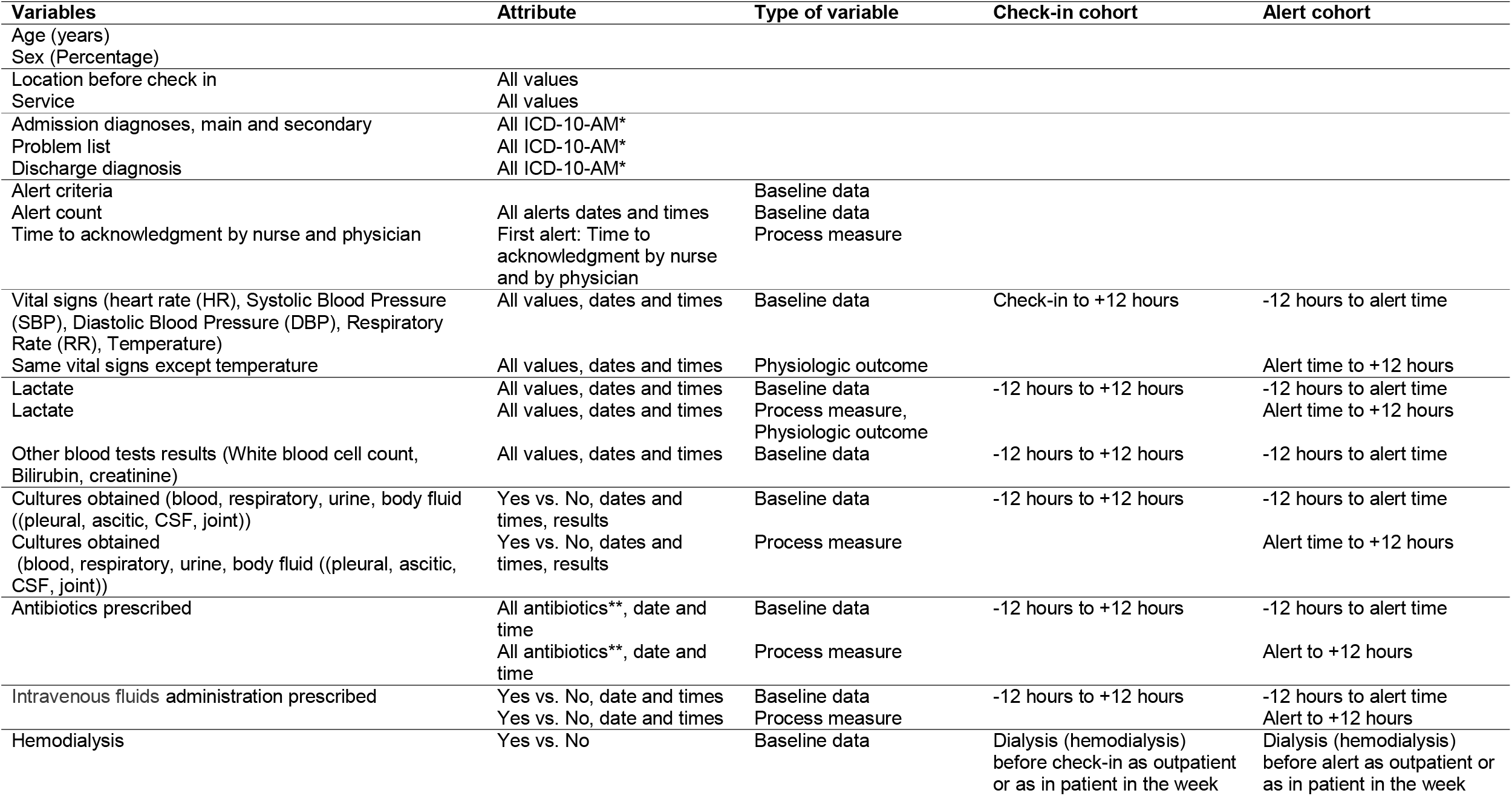
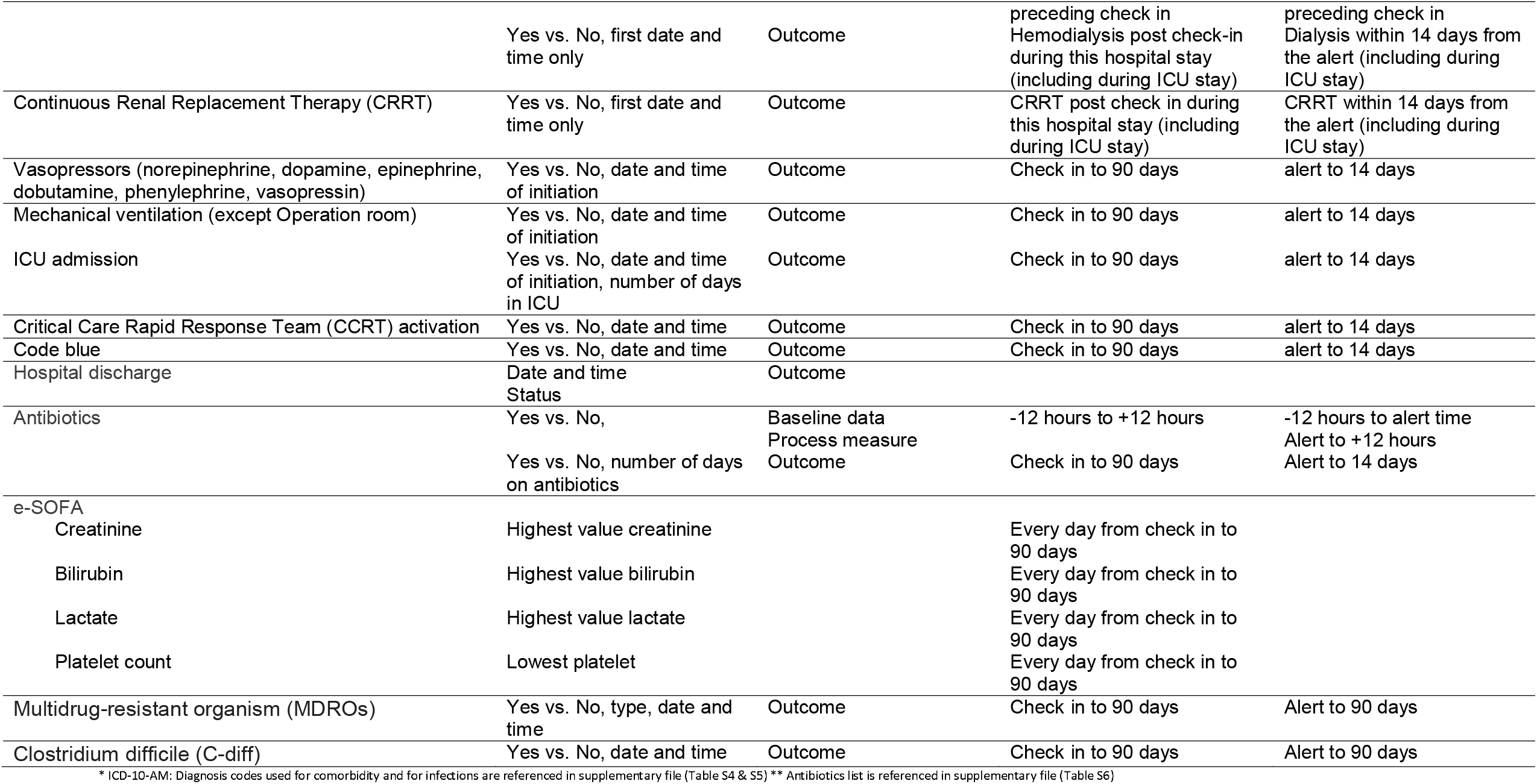
Time windows for data extraction. All data points within the time window will be extracted.

### Baseline Data (ITT and alert cohorts-Supplementary file Table S7)

- Age and Sex
- Admission source: emergency room, operating room, clinic, ICU, others
- Admitting ward: medical, surgical, oncology, mixed (any combination)
- Co morbidities: end-stage renal disease, cancer without metastasis, immune-compromised non-complicated, diabetes, complicated diabetes, congestive heart failure, acquired immunodeficiency syndrome, moderate to severe chronic kidney disease, myocardial infarction, chronic pulmonary disease, peripheral vascular disease, stroke or transient ischemic attack, dementia, hemiplegia or paraplegia, connective tissue disease, peptic ulcer disease, mild liver disease, moderate to severe liver disease
- Charlson Comorbidity Index
- Source of infection on admission to the ward: No infection, pneumonia, urinary tract infection, skin and soft tissue infection, intra-abdominal infection, other infections
- Dialysis
- Alert count, time to first alert and the parameters leading to alert (respiratory rate ≥22 breath/minute, systolic blood pressure ≤100 mmHg, GCS <15)

### Physiological parameters and treatments at baseline (ITT population) and pre-alert (Alert population) (Supplementary file Table S8)

- Systolic and diastolic blood pressure: lowest values in the first 12 hours of check in to the ward (ITT population) and in the 12 hours before the alert
- Heart rate and respiratory rate: highest values in the first 12 hours of check in to the ward (ITT population) and in the 12 hours before the alert
- Temperature: highest value and lowest value in the first 12 hours of check in to the ward (ITT population) and in the 12 hours before the alert
- Lactate level: highest lactate 12 hours before check in to the ward to 12 hours of check-in to the ward (ITT population) and highest value in the 12 hours before the alert (alert population).
- White blood cell count, bilirubin, and creatinine: highest values in the 12 hours before check-in to the ward to 12 hours after check-in to the ward (ITT population) and highest value in the 12 hours before alert (alert population)
- Blood, respiratory, urine and body fluid cultures: percentage of patients with cultures ordered in the 12 hours before check-in to the ward to 12 hours after check-in to the ward (ITT population) and in the 12 hours before alert (alert population)
- Intravenous fluid (NS, 1/2NS, D5NS, D51/2NS, LR, D5LR, albumin 5%, 20%) given in the 12 hours before check-in to the ward to 12 hours after check-in to the ward (ITT population) and in the 12 hours before the alert (alert population)
- Antibiotics: percentage of patients on antibiotics in the 12 hours before check-in to the ward to 12 hours after check-in to the ward (ITT population) and in the 12 hours before alert (alert population)

### Processes measures (Supplementary file Table S9)

- Post-alert lactate level
  ▪ Percentage of patients with lactate reported 12 hours if not reported in the 12 hours before alert
  ▪ Highest value reported in the 12 hours after the alert
- Post-alert blood culture:
  ▪ Percentage of patients with blood culture ordered in 12 hours if not performed in the 12 hours before alert
- Post-alert respiratory, urine and body fluid cultures
  ▪ Percentage of patients with respiratory, urine and body fluid cultures ordered in 12 hours if not performed in the 12 hours before alert
  ▪ Intravenous fluid administered in 12 hours after alert (yes, no)
- Post-alert antibiotics
  ▪ Percentage of patients who were not on antibiotics in the 12 hours before alert and had new antibiotic administered within 3 and 12 hours of alert
  ▪ Percentage of patients who were on antibiotics in the 12 hours before alert and had new antibiotic administered within 3 and 12 hours of alert
- Post-alert systolic blood pressure: lowest value in the 12 hours after the alert.
- Post-alert diastolic blood pressure: lowest value in the 12 hours after the alert
- Post-alert heart rate: highest value in the 12 hours after the alert
- Post-alert respiratory rate: highest value respiratory in the 12 hours after the alert

### Outcomes

#### Primary outcome (Supplementary file Table S10)

In-hospital mortality, percentage of patients who die in the hospital up to 90 days

#### Secondary outcomes

- Outcome measures
  ▪ Hospital length of stay (LOS), censored at 90 days
  ▪ Transfer to ICU within 90 days (ITT cohort) and 14 days of alert (alert cohort)
  ▪ ICU-free days in the first 90 days (ITT cohort and alert cohort)
  ▪ Critical Care Response team (CCRT) activation within 90 days (ITT cohort) and 14 days of alert (alert cohort)
  ▪ Cardiac arrest within 90 days (ITT cohort) and 14 days of alert (alert cohort)
  ▪ The need for mechanical ventilation, vasopressor therapy, incident renal replacement therapy within 90 days (ITT cohort) and 14 days of alert (alert cohort)
  ▪ Organ dysfunction during hospitalization up to 90 days, assessed by electronic SOFA (32)
- Balancing measures
  ▪ Antibiotic-free days within 90 days (ITT cohort and alert cohort)
  ▪ The acquisition of multidrug resistant organisms within 90 days (ITT cohort and alert cohort)
  ▪ Clostridium difficile infection within 90 days (ITT cohort and alert cohort

### Sample size

The sample size for this stepped-wedge cluster RCT [(33-37)] was calculated for 45 clusters with 10 time periods (1 baseline step period followed by 9 steps) using Power Analysis and Sample Size software (PASS 15 Power Analysis and Sample Size Software (2017)). NCSS, LLC. Kaysville, Utah, USA, ncss.com/software/pass). At each of the 9 steps, 5 clusters (wards) will switch from masked alert to active alert. We calculated a baseline rate of 3.13% for mortality in the hospital by day 90 in the “Possible Sepsis Alert” cohort from historical data obtained from the hospital information system. Based on the same dataset, 18.3% of eligible ward patients had sepsis alert with hospital mortality of 8.2% compared to 2.0% in the non-alert patients. For sample size calculations, we made the following assumptions: A) the impact on mortality of implementing the alert occurs only in patients with the alert, B) Only half of patients with alert have sepsis, C) 90% of deaths among the patients with qSOFA occurred among septic patients, D) early intervention resulting from the sepsis alert will reduce the sepsis-related mortality by 50%, resulting in reduction from 8.2% to 4.1% in the patients with the alert and would lead to an overall change in hospital mortality for the whole cohort from 3.13% to 2.46%, risk difference of -0.67%.), E) 80 % power using two-sided Wald Z-Test and significance level of 5% and, F) an intra-cluster correlation (ICC, a measure of the relatedness of cluster) of 0.22 as estimated from retrospective electronic database. As such, a risk difference of mortality in hospital by 90 days by -0.67% (from 3.13% to 2.46%) requires a total sample size of 62550 subjects (average of 1390 subjects per cluster with an average of 139 subjects per cluster per time period). With all five hospitals combined, this is expected to require 20 months (2 months per time period).

### Statistical analysis

For the SCREEN trial, the analysis will be implemented under the guiding principles of the ICH E9 for the analysis of randomized controlled trials.(38) The stepped wedge design is essentially a matched design with before and after comparisons for each unit of randomization. In this case, a unit is a ward where each ward is randomized to a particular starting point.

Raw data will be processed in accordance with the best practices for raw data management to identify any inaccuracies or incompleteness in advance of the statistical analysis. In order to accomplish this task, all interval variables will be checked and summarized in terms of maximum and minimum values. Minimum and maximum values will be checked and compared against the nominal maximum and minimum value of each variable, and variables with implausible values will be flagged. All variables will be summarized and reported for the study using descriptive statistics. Interval variables will be summarized and reported in terms of n, mean and median, standard deviation, the first and third quartiles. Categorical variables will be summarized and reported in terms of frequency distribution. All demographic and clinical variables would be summarized between study groups at baseline by wards and overall.

### Analysis of the primary endpoint

The analysis for the primary outcome will be conducted on the ITT cohort. The rate of all-cause hospital mortality by day 90 will be compared between patients in the intervention wards and control wards using Poisson mixed effect model by adding an offset term to account for variation due to lengths of time to the control and intervention periods. The clusters would be considered as random effect and each step as fixed effect in the model.

### Analysis of secondary endpoints

For transfer to ICU, the proportion of patients transferred to ICU will be compared between the sepsis alert and the no sepsis no alert group using Chi-squared tests, with adjustment for cluster sampling. Possible confounding will be assessed using mixed-effects logistic regression models with random effect for cluster and fixed effect for each step. All covariates that were significantly associated (Chi-squared test adjusted for cluster sampling) with the outcome (proportion of patient transfer to ICU) will be included in the final model. The other secondary endpoints CCRT activation, cardiac arrests, mechanical ventilation, vasopressor therapy, renal replacement therapy and hospital mortality by day 180 will be analyzed in a similar fashion as described above for transfer-to-ICU endpoint.

For hospital length of stay (LOS), the number of days patient stayed at the hospital will be analyzed using negative binomial mixed model with random effect for cluster and fixed effect for each step.

### Subgroups analysis

We will perform the following a priori subgroup analyses: medical, surgical and oncology wards, age > and ≤ 65 years, patients already on antibiotics versus those not on antibiotics at the time of positive qSOFA, patients with alerts within 48 hours of check-in to ward versus patients with later alerts and patients in the different hospitals.

All analyses will be conducted with SAS version 9.4. There will be no interim analysis due to the nature of implementation.

### Analysis of the Alert cohort

Secondary analyses of the alert cohort will follow the same approach used in the ITT cohort.

### Ethical considerations

This is a quality improvement project that is implemented in the participating hospitals in phases. The stepped wedge design is a well-accepted for this type of project,(39) since it ensures that all patients will eventually receive an intervention that is considered to be beneficial. It is expected that the quality of care might improve with time by using the electronic sepsis alert and detecting all at high risk patients. Moreover, the study does not interfere with the routine management of patients and does not require any direct interaction between the research team and patients.(40). Additionally, data are obtained from the electronic medical records for a large sample size of ward patients. Therefore, this study is considered as minimal risk and consent is not required, similar to other studies of the same nature.(41, 42)

## Discussion

This trial evaluates the value of electronic screening for sepsis in patients hospitalized in the wards. Screening for sepsis is of substantial benefit as sepsis is frequent in hospitalized patients, is often under-recognized, is a common cause of organ dysfunction and death, and earlier appropriate management is associated with improved outcomes.(3-6) It is estimated that 32% of sepsis cases are identified in hospital wards,(43) such that hospital-onset sepsis complicates 1 in 200 hospitalizations with an associated mortality of > 30%.(44) (45) Sepsis management is often delayed among patients hospitalized in the ward.(46) (47) A study found that lactate levels were measured within the mandated window in only 32% of patients with severe sepsis in the ward compared with 55% in the ICU and 79% in the ED.(47) Delayed lactate level measurement was associated with longer time to antibiotic administration.(47) The evidence for electronic alerts to early recognize sepsis and improve care processes and outcomes in general ward patients is modest,(48) which affirms the need for an RCT. The stepped-wedge approach to include wards in this trial avoids several pitfalls and retains controlled data elements and randomization.

The optimal tool for screening for sepsis has been debated. A qSOFA score ≥ 2 has been suggested for identification of patients at risk of sepsis outside the ICU. A study in medical wards (481 patients) found that qSOFA had lower sensitivity (44.7% vs. 80.0%), but higher specificity (83.6% vs. 25.7%) and positive predictive value (75.5% vs. 54.8%) for predicting sepsis compared with SIRS.(49) A large prospective cohort study of approximately 1 million hospitalized patients in 85 US hospitals found that 27.0% were qSOFA-positive within 1 day of admission.(50) The sensitivities of qSOFA for suspected infection and sepsis were 41% and 63%, respectively and the positive predictive values were 31% and 17%, respectively.(50) As we prepared for this trial, we initially used SIRS to build the electronic screening tool. Based on analysis of historical data from our hospital information system, qSOFA identified ward patients who were at risk for subsequent ICU admission and death more frequently and earlier than SIRS.(51) Therefore, we revised our alert system to be based on qSOFA. Similar results were observed in multiple systematic reviews.(52-55) SIRS was found to be more sensitive than qSOFA for the diagnosis of sepsis (risk ratio, 1.32; 95% confidence interval, 0.40-2.24; *I* ^2^ = 100%).(52) The pooled specificity of qSOFA in patients with suspected infection outside the ICU was 79.6% (95% confidence interval, 73.3-84.7%).(53) However, current evidence favored qSOFA over SIRS as a predictor of hospital mortality (risk ratio, 0.03; 95% confidence interval, 0.01-0.05; *I* ^2^ = 48%).(52) Therefore, qSOFA may be better than SIRS for identifying high-risk patients.

Real-time electronic screening for sepsis using the electronic record provides multiple advantages over other methods, as it is considered a low-cost solution for more reliable, reproducible, unbiased, and sustainable screening in large hospitals.(56) However, the challenges encountered with our electronic alert development and implementation include resource allocation, changing and un-agreed-upon sepsis screening tools (qSOFA and SIRS), charting behaviors, alert fatigue (false positive), inappropriate response (false negative) and differences in health care delivery models.(57) (58) We have conducted multiple presentations to ward medical and nursing staff and provided online materials on the alert system and sepsis management in line with the Surviving Sepsis Campaign guidelines.(7, 31) The actual management of patients with positive sepsis alert is left to the discretion of the treating team. We also utilized dashboards during the study to provide opportunities for ward medical and nursing leaders to review and improve provided care.

The SCREEN trial provides an opportunity for a novel trial design and analysis of routinely collected and entered data to evaluate the effectiveness of an intervention (sepsis alert) for a common medical problem (sepsis in ward patients). Its results may open the door for other trials on other interventions in other conditions.

## Trial status

The trial started in October 1, 2019 and is expected to continue for 20 months enrolling more than 62550 hospitalized patients.

This protocol is the 3^rd^ version dated September 15, 2019.

## Data Availability

The datasets will be available from the corresponding author as per the regulations of KAIMRC.

## Abbreviations

CCRT: Critical Care Response Team
ED: Emergency department
EMR: Electronic Medical Record
ICU: Intensive Care Unit
ITT: Intention To Treat
LOS: Length of Stay
qSOFA: Quick Sequential Organ Failure Assessment
SIRS: Systemic Inflammatory Response Syndrome
SOFA: Sequential Organ Failure Assessment

## Declarations

### Ethics approval and consent to participate

The study was approved by the National Guard Health Affairs Institutional Review Board (IRB). Informed consent was waived by the IRB because of the nature of the study.

### Consent for publication

Not applicable.

### Availability of data and materials

The datasets will be available from the corresponding author as per the regulations of KAIMRC.

### Authors’ contributions

YA is the Chief Investigator; conception and design, analytical plan, drafting of the manuscript, critical revision of the manuscript for important and intellectual content. AS, AM, MZ, AAQ, AS, AK, FR, HG, EG, RH, EQ, NT, HD, RV, SA, MH, ZY, SS, FH, KA, MK, AQ, MA, AA, NS, AHA, AY, WJ, JA, SQ, AH contributed to the development of the protocol, critical revision of the manuscript for important intellectual content. All authors read and approved the final manuscript.

## SCREEN Trial Group

**Ahmed Al Arfaj, MD**

College of Medicine, King Saud bin Abdulaziz University for Health Sciences, King Abdullah International Medical Research Center, Al Ahsa, Saudi Arabia Ministry of National Guard Health Affairs, Eastern Region, Saudi Arabia

**Mohamed S. Al Moammary, ABIM FRCP(Edin) MBA-LS FCCP**

College of Medicine, King Saud bin Abdulaziz University for Health Sciences, King Abdullah International Medical Research Center, Riyadh Saudi Arabia King Abdulaziz Medical City Ministry of National Guard Health Affairs, Riyadh, Saudi Arabia

**Soud Rasheed, MBBS, FRCPC**

College of Medicine, King Saud Bin Abdulaziz University for Health Sciences King Abdullah International Medical Research Center King Abdulaziz Medical City Ministry of National Guard Health Affairs, Riyadh, Saudi Arabia

**Turki Alwasaidi, MD**

College of Medicine, King Saud Bin Abdulaziz University for Health Sciences King Abdullah International Medical Research Center Prince Mohammed bin Abdulaziz Hospital Ministry of National Guard Health Affairs, Madinah, Saudi Arabia

**Amal Matroud, RN**

King Saud Bin Abdulaziz University for Health Sciences King Abdullah International Medical Research Center Nursing Services Department, King Abdulaziz Medical City Ministry of National Guard Health Affairs, Riyadh, Saudi Arabia

**Amar M Alhasani, MD**

College of Medicine, King Saud Bin Abdulaziz University for Health Sciences King Abdullah International Medical Research Center Prince Mohammed bin Abdulaziz Hospital Ministry of National Guard Health Affairs, Madinah, Saudi Arabia

**Haifa Al Shammari, RN**

College of Medicine, King Saud Bin Abdulaziz University for Health Sciences King Abdullah International Medical Research Center Intensive Care Department, King Abdulaziz Medical City Ministry of National Guard Health Affairs, Riyadh, Saudi Arabia

**Majid M Alshamrani, MD**

College of Medicine, King Saud Bin Abdulaziz University for Health Sciences King Abdullah International Medical Research Center Infection Prevention and Control Department, King Abdulaziz Medical City Ministry of National Guard Health Affairs, Riyadh, Saudi Arabia

**Saleh Qasim, MD**

College of Medicine, King Saud Bin Abdulaziz University for Health Sciences King Abdullah International Medical Research Center Intensive Care Department, King Abdulaziz Medical City Ministry of National Guard Health Affairs, Riyadh, Saudi Arabia

**Saeed Obbed, MD**

College of Medicine, King Saud Bin Abdulaziz University for Health Sciences King Abdullah International Medical Research Center Intensive Care Department, King Abdulaziz Medical City Ministry of National Guard Health Affairs, Riyadh, Saudi Arabia

**Adnan A Munshi**

King Saud bin Abdulaziz University for Health Sciences, King Abdullah International Medical Research Center Medical Imaging Informatics, King Abdulaziz Medical City Ministry of National Guard Health Affairs, Jeddah, Saudi Arabia

**Hadia Al Tabsh, RN, MSN**

King Saud Bin Abdulaziz University for Health Sciences King Abdullah International Medical Research Center Nursing Services Department, King Abdulaziz Medical City Ministry of National Guard Health Affairs, Jeddah, Saudi Arabia

**Basem R Banat, RN**

King Saud Bin Abdulaziz University for Health Sciences King Abdullah International Medical Research Center Nursing Services Department, King Abdulaziz Medical City Ministry of National Guard Health Affairs, Jeddah, Saudi Arabia

**Omar Abuskout, RN, MSN**

King Saud Bin Abdulaziz University for Health Sciences King Abdullah International Medical Research Center Nursing Services Department, King Abdulaziz Medical City Ministry of National Guard Health Affairs, Jeddah, Saudi Arabia

**Anna Liza Marcelo, RN**

King Saud Bin Abdulaziz University for Health Sciences King Abdullah International Medical Research Center Nursing Services Department, King Abdulaziz Medical City Ministry of National Guard Health Affairs, Jeddah, Saudi Arabia

**Mayadah M Alhabshi, RN, DNE, MSN**

King Saud Bin Abdulaziz University for Health Sciences King Abdullah International Medical Research Center Quality and Patient Safety Department, King Abdulaziz Medical City Ministry of National Guard Health Affairs, Jeddah, Saudi Arabia

**Ibrahim J. Jaber, RN**

King Saud Bin Abdulaziz University for Health Sciences King Abdullah International Medical Research Center Nursing Services Department, Prince Mohammed bin Abdulaziz Hospital Ministry of National Guard Health Affairs, Madinah, Saudi Arabia

**Mohammad Shahin, RN, MSN**

King Saud Bin Abdulaziz University for Health Sciences King Abdullah International Medical Research Center Nursing Services Department, King Abdulaziz Hospital Ministry of National Guard Health Affairs, Al Ahsa, Saudi Arabia

**Jamielah Yaakob, RN**

King Saud Bin Abdulaziz University for Health Sciences King Abdullah International Medical Research Center Nursing Services Department, King Abdulaziz Hospital Ministry of National Guard Health Affairs, Al Ahsa, Saudi Arabia

**Hanan Al Somali, RN**

College of Medicine, King Saud Bin Abdulaziz University for Health Sciences King Abdullah International Medical Research Center Nursing Services Department, Imam Abdulrahman Al Faisal Hospital Ministry of National Guard Health Affairs, Dammam, Saudi Arabia

**Clara Masala, RN**

College of Medicine, King Saud Bin Abdulaziz University for Health Sciences King Abdullah International Medical Research Center Nursing Services Department, Imam Abdulrahman Al Faisal Hospital Ministry of National Guard Health Affairs, Dammam, Saudi Arabia

**Mohammed Al Qarni, MHHA, RRT**

College of Medicine, King Saud Bin Abdulaziz University for Health Sciences King Abdullah International Medical Research Center Quality and Patient Safety Department, King Abdulaziz Medical City Ministry of National Guard Health Affairs, Riyadh, Saudi Arabia

**Jamal Chalabi, MD**

College of Medicine, King Saud Bin Abdulaziz University for Health Sciences King Abdullah International Medical Research Center Intensive Care Department, King Abdulaziz Hospital Ministry of National Guard Health Affairs, Al Ahsa, Saudi Arabia

**Johanna E Greyvenstein, RN, MSN**

King Saud Bin Abdulaziz University for Health Sciences King Abdullah International Medical Research Center Nursing Services Department, King Abdulaziz Hospital Ministry of National Guard Health Affairs, Al Ahsa, Saudi Arabia

**Abdul Rahman Jazieh, MD, MPH**

College of Medicine, King Saud Bin Abdulaziz University for Health Sciences King Abdullah International Medical Research Center Oncology Department, King Abdulaziz Medical City Riyadh, Saudi Arabia

**Noha Omaish, RN**

King Saud Bin Abdulaziz University for Health Sciences King Abdullah International Medical Research Center Nursing Services Department, Prince Mohammed bin Abdulaziz Hospital Ministry of National Guard Health Affairs, Madinah, Saudi Arabia

**Azura Abdrahim, RN**

King Saud Bin Abdulaziz University for Health Sciences King Abdullah International Medical Research Center Nursing Services Department, Prince Mohammed bin Abdulaziz Hospital Ministry of National Guard Health Affairs, Madinah, Saudi Arabia

**Mohammad Abdrabo, MD**

College of Medicine, King Saud Bin Abdulaziz University for Health Sciences King Abdullah International Medical Research Center Quality and Patient Safety Department, Prince Mohammed bin Abdulaziz Hospital Ministry of National Guard Health Affairs, Madinah, Saudi Arabia

**Abdullah Al Hamdan**

King Saud bin Abdulaziz University for Health Sciences King Abdullah International Medical Research Center Medical Imaging Informatics, King Abdulaziz Medical City Ministry of National Guard Health Affairs, Jeddah, Saudi Arabia

**Abdulaziz Al Qasem**

King Saud bin Abdulaziz University for Health Sciences King Abdullah International Medical Research Center Information Systems and Informatics Division, King Abdulaziz Hospital Ministry of National Guard Health Affairs, Al Ahsa, Saudi Arabia

**Hattan Esilan, MD**

King Saud bin Abdulaziz University for Health Sciences King Abdullah International Medical Research Center Quality and Patient Safety Department, King Abdulaziz Hospital Ministry of National Guard Health Affairs, Al Ahsa, Saudi Arabia

