## Supplementary file for "Electronic early notification of sepsis in hospitalized ward patients: a study protocol for a stepped-wedge cluster randomized controlled trial"

### **Table S1:** Total number of eligible wards in the participating hospitals**.**

| **Center** | **Number of wards** |
| --- | --- |
| King Abdulaziz Medical City – Riyadh | 25 wards |
| King Abdulaziz Medical City – Jeddah | 9 wards |
| King Abdulaziz Hospital – Al Ahsa | 6 wards |
| Prince Mohammad Bin Abdulaziz Hospital – Madinah | 4 wards |
| Imam Abdulrahman Al Faisal Hospital – Dammam | 2 wards |
| **Total** | **46*** |

* Two of the wards were combined for purposes of randomization

**Table S2:** Definitions used in the SCREEN trial.

| **Item** | **Description** |
| --- | --- |
| **Time 0** | Date and time when the patient is checked in for the first time to the ward. If transferred to different wards, take the first date and time only. |
| **Alert time** | When the patient meets the alert criteria; for patients who have multiple alerts, we will take only the first alert |
| **Observation time** | The whole hospital stay from time of check-in to the ward to time of discharge from the hospital censored at 90 days. The observation period excludes stays in the operating room, daycare operation room, operation cardiac, outpatient, post-anesthesia care unit, dental outpatient, day surgery, daycare outpatient, intensive care unit, trauma intensive care unit, surgical intensive care unit, Intermediate care unit, progressive care unit, medical cardiac intensive care unit, neurointensive care unit, burn unit). |
| **Alert population** | Patients who had one alert or more during hospitalization. However, detailed data will be included only for the first alert. For subsequent alerts, only alert count will be provided. |

**Table S3:** Guidelines for nurses and physicians.

**GUIDELINES FOR PHYSICIANS**

*What to do if my patient has an alert?*

1- A sign will appear in BESTCare on patient list page when a patient meets the qSOFA criteria “Possible Sepsis Alert” in BESTCare patients’ list

“Possible Sepsis” alert sign

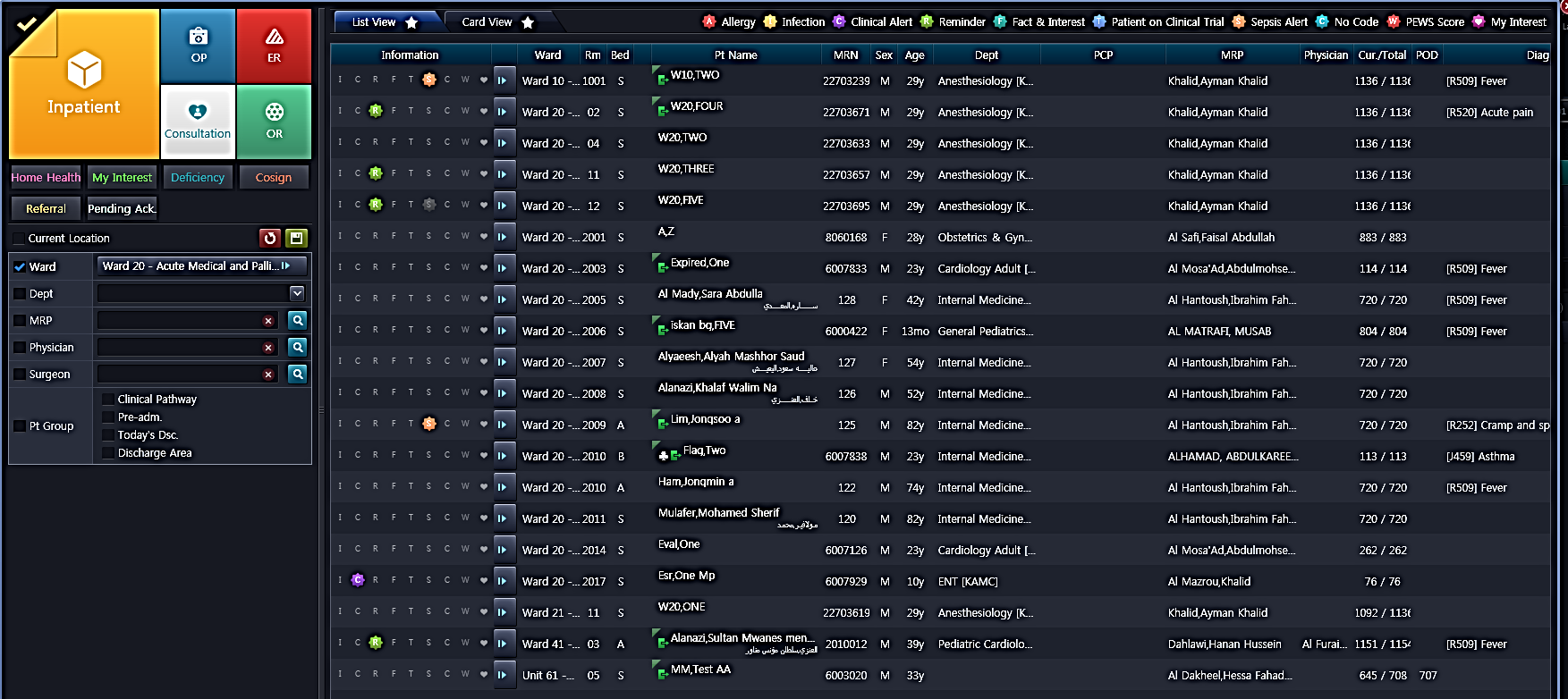

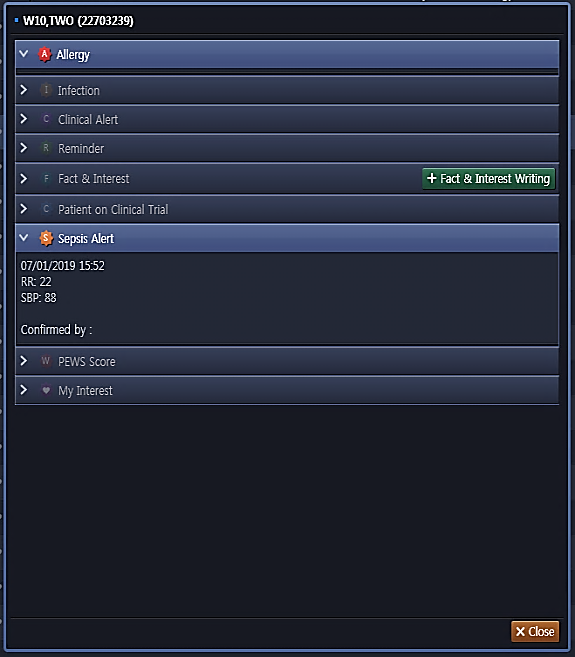

If you click on the alert sign:

2- A pop-up message will appear in the patient record when patient file is opened. The message asks the physician to assess the patient and document if the patient has sepsis.

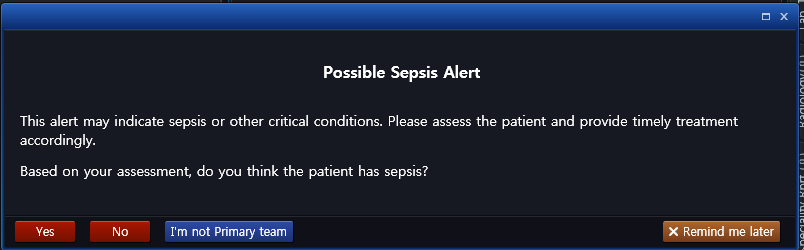

**GUIDELINES FOR NURSES**

1- Each nurse will have a card that shows who meets qSOFA criteria and when an alert will be activated as shown below.

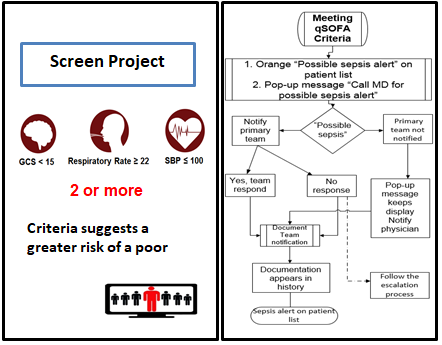

2-
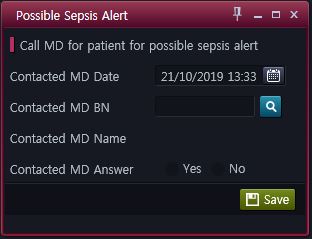
The bedside nurse will receive a message when a patient meets qSOFA criteria. The message asks the nurse to immediately inform the primary team and to document that the primary team has been notified. In case of nor response, the nurse will follow an escalation process.

3- The charge nurse will be alerted that the patient met the criteria through an iPod application.

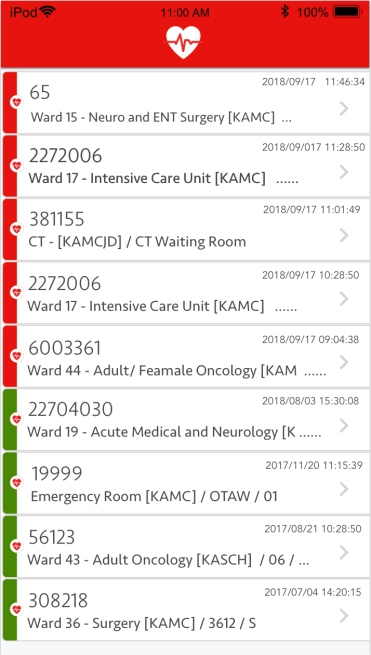

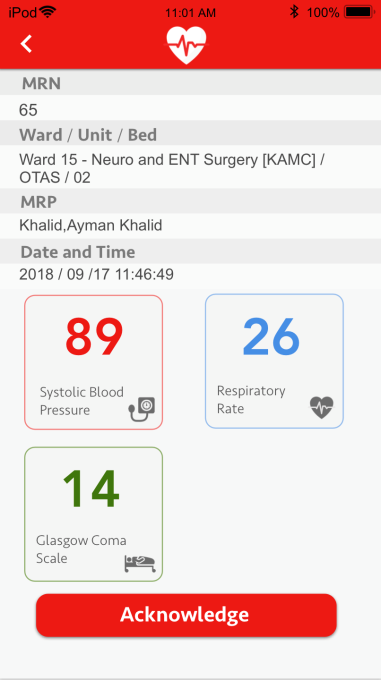

**Table S4:** The ICD-10-AM codes for comorbid conditions.

| **Comorbidity** | **ICD-10-AM Codes** |
| --- | --- |
| Myocardial infarction | I21.x, I22.x, I25.2 |
| Congestive heart failure | I09.9, I11.0, I13.0, I13.2, I25.5, I42.0, I42.5 - I42.9, I43.x, I50.x, P29.0 |
| Peripheral vascular disease | I70.x, I71.x, I73.1, I73.8, I73.9, I77.1, I79.0, I79.2, K55.1, K55.8, K55.9, Z95.8, Z95.9 |
| Cerebrovascular disease | G45.x, G46.x, H34.0, I60.x - I69.x |
| Dementia | F00.x - F03.x, F05.1, G30.x, G31.1 |
| Chronic pulmonary disease | I27.8, I27.9, J40.x - J47.x, J60.x - J67.x, J68.4, J70.1, J70.3 |
| Rheumatic disease | M05.x, M06.x, M31.5, M32.x - M34.x, M35.1, M35.3, M36.0 |
| Peptic ulcer disease | K25.x - K28.x |
| Mild liver disease | B18.x, K70.0 - K70.3, K70.9, K71.3 - K71.5, K71.7, K73.x, K74.x, K76.0, K76.2 - K76.4, K76.8, K76.9, Z94.4 |
| Diabetes without chronic complication | E10.0, E10.1, E10.6, E10.8, E10.9, E11.0, E11.1, E11.6, E11.8, E11.9, E12.0, E12.1, E12.6, E12.8, E12.9, E13.0, E13.1, E13.6, E13.8, E13.9, E14.0, E14.1, E14.6, E14.8, E14.9 |
| Diabetes with chronic complication | E10.2 - E10.5, E10.7, E11.2 - E11.5, E11.7, E12.2 - E12.5, E12.7, E13.2 - E13.5, E13.7, E14.2 - E14.5, E14.7 |
| Hemiplegia or paraplegia | G04.1, G11.4, G80.1, G80.2, G81.x, G82.x, G83.0 - G83.4, G83.9 |
| Renal disease | I12.0, I13.1, N03.2 - N03.7, N05.2 - N05.7, N18.x, N19.x, N25.0, Z49.0 - Z49.2, Z94.0, Z99.2 |
| Any malignancy, including lymphoma and leukemia, except malignant neoplasm of skin | C00.x - C26.x, C30.x - C34.x, C37.x - C41.x, C43.x, C45.x - C58.x, C60.x - C76.x, C81.x - C85.x, C88.x, C90.x - C97.x |
| Moderate or severe liver disease | I85.0, I85.9, I86.4, I98.2, K70.4, K71.1, K72.1, K72.9, K76.5, K76.6, K76.7 |
| Metastatic solid tumor | C77.x - C80.x |
| AIDS/HIV | B20.x - B22.x, B24.x |

**Table S5:** The ICD-10-AM codes used for infectious diseases.

| **Infectious Disease Group** | **ICD-10-AM Code** |
| --- | --- |
| Acute lower respiratory tract infections | A481, A482, B59, J09, J100, J101, J108, J110, J111, J118, J120, J121, J122, J128, J129, J13, J14, J150, J151, J152, J153, J154, J155, J156, J157, J158, J159, J160, J168, J170, J171, J172, J173, J178, J180, J181, J182, J188, J189, J200, J201, J202, J203, J204, J205, J206, J207, J208, J209, J210, J218, J219, J22 |
| Breast infections | N61 |
| Central nervous system general infections | G000, G001, G002, G003, G008, G009, G01, G020, G021, G028, G030, G039, G040, G041, G042, G048, G049, G050, G051, G052, G058, G060, G061, G062, G07, G08, G09, G610 |
| Central nervous system viral infections | A801, A802, A803, A804, A809, A811, A812, A818, A819, A820, A821, A829, A830, A831, A832, A833, A834, A835, A836, A838, A839, A840, A841, A848, A849, A850, A851, A852, A858, A86, A870, A871, A872, A878, A879, A880, A881, A888, A89 |
| Connective tissue infectious | M0210, M0211, M0212, M0213, M0214, M0215, M0216, M0217, M0218, M0219, M0230, M0231, M0232, M0233, M0234, M0235, M0236, M0237, M0238, M0239, M0300, M0301, M0302, M0303, M0304, M0305, M0306, M0307, M0308, M0309, M0310, M0311, M0312, M0313, M0314, M0315, M0316, M0317, M0318, M0319, M0320, M0321, M0322, M0323, M0324, M0325, M0326, M0327, M0328, M0329, M0360, M0361, M0362, M0363, M0364, M0365, M0366, M0367, M0368, M0369, M6000, M6001, M6002, M6003, M6004, M6005, M6006, M6007, M6008, M6009, M6300, M6301, M6302, M6303, M6304, M6305, M6306, M6307, M6308, M6309, M6310, M6311, M6312, M6313, M6314, M6315, M6316, M6317, M6318, M6319, M6320, M6321, M6322, M6323, M6324, M6325, M6326, M6327, M6328, M6329, M6500, M6501, M6502, M6503, M6504, M6505, M6506, M6507, M6508, M6509, M6510, M6511, M6512, M6513, M6514, M6515, M6516, M6517, M6518, M6519, M6800, M6801, M6802, M6803, M6804, M6805, M6806, M6807, M6808, M6809, M7100, M7101, M7102, M7103, M7104, M7105, M7106, M7107, M7108, M7109, M7110, M7111, M7112, M7113, M7114, M7115, M7116, M7117, M7118, M7119, M8960, M8961, M8962, M8963, M8964, M8965, M8966, M8967, M8968, M8969 |
| Enteric infections | A000, A001, A009, A010, A011, A012, A013, A014, A020, A021, A022, A028, A029, A030, A031, A032, A033, A038, A039, A040, A041, A042, A043, A044, A045, A046, A047, A048, A049, A050, A051, A052, A053, A054, A058, A059, A060, A061, A062, A063, A064, A065, A066, A067, A068, A069, A070, A071, A072, A073, A078, A079, A080, A081, A082, A083, A084, A085 |
| Enteric symptoms | A090, A099, I880, K528, K529, R11 |
| Gastrointestinal tract infections | K230, K231, K250, K251, K252, K253, K254, K255, K256, K257, K259, K260, K261, K262, K263, K264, K265, K266, K267, K269, K270, K271, K272, K273, K274, K275, K276, K277, K279, K280, K281, K282, K283, K284, K285, K286, K287, K289, K293, K294, K295, K350, K351, K359, K36, K37, K610, K611, K612, K613, K614, K630, K632, K650, K678, K908 |
| Heart and circulatory infections | B332, I00, I010, I011, I012, I018, I019, I020, I029, I050, I051, I052, I058, I059, I060, I061, I062, I068, I069, I070, I071, I072, I078, I079, I080, I081, I082, I083, I088, I089, I090, I091, I092, I098, I099, I301, I330, I339, I38, I390, I391, I392, I393, I394, I398, I400, I410, I411, I412, I430, I716, I790, I791 |

| **Infectious Disease Group** | **ICD-10-AM Code** |
| --- | --- |
| Hepatic infections | K750, K770, K830 |
| Joint infections | M0000, M0001, M0002, M0003, M0004, M0005, M0006, M0007, M0008, M0009, M0010, M0011, M0012, M0013, M0014, M0015, M0016, M0017, M0018, M0019, M0020, M0021, M0022, M0023, M0024, M0025, M0026, M0027, M0028, M0029, M0080, M0081, M0082, M0083, M0084, M0085, M0086, M0087, M0088, M0089, M0090, M0091, M0092, M0093, M0094, M0095, M0096, M0097, M0098, M0099, M0100, M0101, M0102, M0103, M0104, M0105, M0106, M0107, M0108, M0109, M0110, M0111, M0112, M0113, M0114, M0115, M0116, M0117, M0118, M0119, M0120, M0121, M0122, M0123, M0124, M0125, M0126, M0127, M0128, M0129, M0130, M0131, M0132, M0133, M0134, M0135, M0136, M0137, M0138, M0139, M0140, M0141, M0142, M0143, M0144, M0145, M0146, M0147, M0148, M0149, M0150, M0151, M0152, M0153, M0154, M0155, M0156, M0157, M0158, M0159, M0160, M0161, M0162, M0163, M0164, M0165, M0166, M0167, M0168, M0169, M0180, M0181, M0182, M0183, M0184, M0185, M0186, M0187, M0188, M0189 |
| Kidney infections | N000, N001, N002, N003, N004, N005, N006, N007, N008, N009, N050, N051, N052, N053, N054, N055, N056, N057, N058, N059, N10, N136, N151 |
| Meningococcal disease | A390, A391, A392, A393, A394, A395, A398, A399 |
| Osteomyelitis | M4620, M4621, M4622, M4623, M4624, M4625, M4626, M4627, M4628, M4629, M4630, M4631, M4632, M4633, M4634, M4635, M4636, M4637, M4638, M4639, M4640, M4641, M4642, M4643, M4644, M4645, M4646, M4647, M4648, M4649, M4650, M4651, M4652, M4653, M4654, M4655, M4656, M4657, M4658, M4659 |
| Other bacterial infections | A200, A201, A202, A203, A207, A208, A209, A210, A211, A212, A213, A217, A218, A219, A220, A221, A222, A227, A228, A229, A230, A231, A232, A233, A238, A239, A240, A241, A242, A243, A244, A250, A251, A259, A260, A267, A268, A269, A270, A278, A279, A280, A281, A282, A288, A289, A300, A301, A302, A303, A304, A305, A308, A309, A310, A311, A318, A319, A320, A321, A327, A328, A329, A33, A34, A35, A360, A361, A362, A363, A368, A369, A370, A371, A378, A379, A38, A420, A421, A422, A427, A428, A429, A430, A431, A438, A439, A440, A441, A448, A449, A480, A483, A484, A488, A490, A491, A492, A493, A498, A499, A65, A660, A661, A662, A663, A664, A665, A666, A667, A668, A669, A670, A671, A672, A673, A679, A680, A681, A689, A690, A691, A692, A698, A699, A70, A710, A711, A719, A740, A748, A749, A750, A751, A752, A753, A759, A770, A771, A772, A773, A778, A779, A78, A790, A791, A798, A799, B950, B951, B952, B953, B9541, B9542, B9548, B955, B956, B957, B958, B960, B961, B962, B9631, B9638, B9639, B964, B965, B966, B967, B9681, B9688 |
| Other infectious diseases | B650, B651, B652, B653, B658, B659, B660, B661, B662, B663, B664, B665, B668, B669, B670, B671, B672, B673, B674, B675, B676, B677, B678, B679, B680, B681, B689, B690, B691, B698, B699, B700, B701, B710, B711, B718, B719, B72, B73, B740, B741, B742, B743, B744, B748, B749, B75, B760, B761, B768, B769, B770, B778, B779, B780, B781, B787, B789, B79, B80, B810, B811, B812, B813, B814, B818, B820, B829, B830, B831, B832, B833, B834, B838, B839, B850, B851, B852, B853, B854, B870, B871, B872, B873, B874, B878, B879, B880, B881, B882, B883, B888, B889, B89, B940, B941, B942, B948, B949, B99, E033, E321, F024, F071, I881, I888, I889, T64 |

| **Infectious Disease Group** | **ICD-10-AM Code** |
| --- | --- |
| Other mycoses | B350, B351, B352, B353, B354, B355, B356, B358, B359, B360, B361, B362, B363, B368, B369, B370, B371, B372, B373, B374, B375, B376, B377, B3781, B3788, B379, B380, B381, B382, B383, B384, B387, B388, B389, B390, B391, B392, B393, B394, B395, B399, B400, B401, B402, B403, B407, B408, B409, B410, B417, B418, B419, B420, B421, B427, B428, B429, B430, B431, B432, B438, B439, B440, B441, B442, B447, B448, B449, B450, B451, B452, B453, B457, B458, B459, B460, B461, B462, B463, B464, B465, B468, B469, B470, B471, B479, B480, B481, B482, B483, B484, B487, B488, B49 |
| Other viral infections | A90, A91, A920, A921, A922, A923, A924, A928, A929, A930, A931, A932, A938, A94, A950, A951, A959, A960, A961, A962, A968, A969, A980, A981, A982, A983, A984, A985, A988, A99, B000, B001, B002, B003, B004, B005, B007, B008, B009, B010, B011, B012, B018, B019, B020, B021, B022, B023, B027, B028, B029, B03, B04, B050, B051, B052, B053, B054, B058, B059, B060, B068, B069, B07, B080, B081, B082, B083, B084, B085, B088, B09, B250, B251, B252, B258, B259, B260, B261, B262, B263, B268, B269, B270, B271, B278, B279, B331, B333, B334, B338, B340, B341, B342, B343, B344, B348, B349, B970, B971, B972, B973, B974, B975, B976, B977, B978 |
| Postoperative infections | T802, T8141, T8142, T826, T827, T835, T836, T845, T846, T847, T8571, T8572, T8578, T874 |
| Reproductive system infections, female | N700, N701, N709, N710, N711, N719, N72, N730, N731, N732, N733, N734, N735, N736, N738, N739, N748, N751, N764, N870, N871, N872, N879 |
| Reproductive system infections, male | N410, N411, N412, N413, N431, N450, N459, N481, N482, N490, N491, N492, N498, N499, N510, N511, N512, N518 |
| Septicaemia | A400, A401, A402, A403, A408, A409, A410, A411, A412, A413, A414, A4150, A4151, A4152, A4158, A418, A419 |
| Skin infections, other | B86, S1013, S1083, S1093, S2013, S2033, S2043, S2083, S3083, S3093, S4083, S5083, S6083, S7083, S8083, S9083, T009, T0903, T1108, T1303, T1403, T633, T634, T793, T8901, T8902 |
| Skin infections, typical | A46, L00, L010, L011, L020, L021, L022, L023, L024, L028, L029, L0301, L0302, L0310, L0311, L032, L033, L038, L039, L040, L041, L042, L043, L048, L049, L050, L080, L081, L088, L089 |
| Tuberculosis | A150, A151, A152, A153, A154, A155, A156, A157, A158, A159, A160, A161, A162, A163, A164, A165, A167, A168, A169, A170, A171, A178, A179, A180, A181, A182, A183, A184, A185, A186, A187, A188, A190, A191, A192, A198, A199, J65, N740, N741 |
| Upper respiratory tract infections | J00, J010, J011, J012, J013, J014, J018, J019, J020, J028, J029, J030, J038, J039, J040, J041, J042, J050, J051, J060, J068, J069, J320, J321, J322, J323, J324, J328, J329, J340, J36, J370, J371, J390, J391 |
| Urinary tract infections | N300, N341, N351, N370, N378, N390 |

**Table S6:** List of antibiotics based on the hospital formulary.

**Antibiotics**

Aztreonam

Azithromycin

Piperacillin/Tazobactam

Vancomycin

Ceftriaxone

Metronidazole

Imipenem/Cilastin

Moxifloxacin

Gentamicin

Meropenem

Clindamycin

Metronidazole

Cefepime

Caspofungin

Fluconazole

Linezolid

Cloxacillin

Amikacin

Cefazolin

Cefuroxime

Colistin

Ciprofloxacin

Ticarcillin/Clavulanate

Ampicillin/Sulbactam

Amoxicillin

Ceftazidime

Ceftazidime/avibactam

Tigecycline

Trimethoprim-sulfamethoxazole

Amphotericin-B

Voriconazole

Anidulafungin

Doxyclycline

**Table S7:** Baseline data.

| **Variable** | **Definitions** |
| --- | --- |
| Age (yr), Median (Q1, Q3) | Age |
| Sex (number, %) | Gender |
| Admission source (number, %) | location of the patient before ward check-in |
| Emergency Room | Emergency department |
| Operating Room | Operative room |
| Clinic | Outpatient |
| Intensive care unit | Intensive care unit (cardiac and non-cardiac) |
| Others | Any other which may include: other hospitals, other ward |
| **Service (number, %)** | |
| Medical | Medical wards include: adult diabetic and endocrine center, Adult Hepatology, Endocrinology & Metabolism; Gastroenterology; Hepatobiliary Sciences; Internal Medicine; Nephrology; Neurology; Palliative Care; Psychiatry; Pulmonology; Rheumatology |
| Surgical | Surgical wards include: adult organ transplant & hepatobiliary surgery, Adult Transplant Nephrology; Anesthesiology, Breast Surgery; General Surgery; Neurosurgery; Oral & Maxillofacial Surgery; Orthopedics; Podiatric Surgery; Plastic Surgery; Thoracic Surgery; Urology Surgery; Vascular Surgery; ENT |
| Hematology-Oncology | Hematology-oncology wards include: Adult hematology; Adult medical oncology; Adult hematology and oncology; Adult Stem Cell Transplant & Cellular Therapy; Radiation Oncology |
| Other | Intensive Care - Adult; Obstetrics & Gynecology; Ophthalmology |
| **Admitting ward (number, %)** | |
| Medical | Riyadh: ward 7, ward 8, ward 10, ward 12, ward 13, ward 19, ward 20, ward 22, ward 23, ward 24, ward 25, AMU  Jeddah: ward 4, ward 5, ward 6  Madinah: ward 8  Dammam: none  Al Ahsa : ward 3, ward 7 |
| Surgical | Riyadh: ward 15, ward 18, ward 36, ward 37, ward 38, ward 39, ward 40, ASU  Jeddah: ward 14, ward 16  Madinah: ward 5  Dammam: none  Al Ahsa: ward 2, ward 8 |
| Oncology | Riyadh: ward 41, ward 43, ward 44  Jeddah: ward 22, ward 23, ward 25  Madinah: none  Dammam: none  Al Ahsa : ward 5 |
| Mixed (any combination) | Riyadh: ward 16 A, B, C, ward 49  Jeddah: ward 3  Madinah: ward 3, ward 6  Dammam: ward 1, ward 3  Al Ahsa: ward 6 |
| **Admission diagnosis, main and** **secondary** | Admission note, problem list, or ICD-10-AM code |

| **Comorbidities (number, %)** |  |
| --- | --- |
| End-Stage Renal Disease (ESRD) | ICD-10-AM * |
| Chronic liver disease |  |
| Cancer without metastasis |  |
| immune-compromised |  |
| Diabetes non-complicated |  |
| Diabetes, complicated |  |
| Congestive heart failure |  |
| Acquired immunodeficiency syndrome |  |
| Moderate to severe Chronic Kidney Disease (CKD) |  |
| Myocardial infarction |  |
| Chronic pulmonary disease |  |
| Peripheral vascular disease |  |
| Stroke or transient ischemic attack |  |
| Dementia |  |
| Hemiplegia or paraplegia |  |
| Connective tissue disease |  |
| Peptic ulcer disease |  |
| Mild liver disease |  |
| Moderate to severe liver disease |  |
| Charlson Comorbidity Index |  |
| **Source of infection on admission (number, %)** |  |
| No infection |  |
| Pneumonia |  |
| Urinary tract infection |  |
| Skin and soft tissue infection |  |
| Intra-abdominal infection  other infections |  |
| **Dialysis** |  |
| Alert count |  |
| Time to first alert |  |
| Alert information criteria leading to triggering (number, %) |  |
| RR≥22 (breath/minute) | Respiratory Rate |
| SBP≤100 mm Hg | Systolic Blood Pressure |
| GCS<15 | Glasgow Coma Scale |

**Table S8:** Physiological parameters and treatments at baseline and pre-alert.

| Variable | Description |
| --- | --- |
| Vital Signs |  |
| Blood Pressure |  |
| Baseline systolic blood pressure | The lowest value in the first 12 hours of check in to the ward |
| Baseline diastolic blood pressure | The lowest value in the first 12 hours of check in to the ward |
| Pre-alert systolic blood pressure | The lowest value in the 12 hours before the alert |
| Pre-alert diastolic blood pressure | The lowest value in the 12 hours before the alert |
| Heart Rate |  |
| Baseline heart rate | The highest value in the first 12 hours of check in to the ward |
| Pre-alert heart rate | The highest value in the 12 hours before the alert |
| Temperature |  |
| Baseline temperature | The highest value and lowest value in the first 12 hours of check in to the ward |
| Pre-alert temperature | The highest value and lowest value in the 12 hours before the alert |
| Respiratory Rate |  |
| Baseline respiratory rate | The highest value in the first 12 hours of check in to the ward |
| Pre-alert respiratory rate | The highest value in the 12 hours before the alert |
| Lab parameters |  |
| Lactate |  |
| Baseline lactate (mmol/L) | The highest value 12 hours before check in to the ward to 12 hours of check-in to the ward |
| Pre-alert lactate (mmol/L) | The highest value in the 12 hours before the alert. |
| Complete Blood Count |  |
| Baseline white blood cells 10^9^/L | The highest value in the 12 hours before check-in to the ward to 12 hours after check-in to the ward |
| Pre-alert white blood cells 10^9^/L | The highest value in the 12 hours before the alert |
| Bilirubin |  |
| Baseline bilirubin (µmol/L) | The highest value in the 12 hours before check-in to the ward to 12 hours after check-in to the ward |
| Pre-alert bilirubin (µmol/L) | The highest value in the 12 hours before the alert |
| Creatinine |  |
| Baseline creatinine (µmol/L) | The highest value in the 12 hours before check-in to the ward to 12 hours after check-in to the ward |
| Pre-alert creatinine µmol/L) | The highest value in the 12 hours before the alert |
| Culture (number, %) |  |
| Baseline blood culture | Blood culture ordered in the 12 hours before check-in to the ward to 12 hours after check-in to the ward, results |
| Pre-alert blood culture | Blood culture ordered in the 12 hours before the alert |
| Baseline respiratory culture | Respiratory culture ordered in the 12 hours before check-in to the ward to 12 hours after check-in to the ward (number, %), results |
| Pre-alert respiratory culture | Respiratory culture ordered in the 12 hours before the alert |
| Baseline urine culture | Urine culture ordered in the 12 hours before check-in to the ward to 12 hours after check-in to the ward, results |
| Pre-alert urine culture | Urine culture ordered in the 12 hours before alert |
| Baseline Body fluid Culture (pleural, ascitic, CSF, joint) | Body fluid culture ordered in the 12 hours before check-in to the ward to 12 hours after check-in to the ward, results |
| Pre-alert Body fluid culture | Body fluid culture ordered in the 12 hours before alert |
| Intravenous fluids (number, %) |  |
| Baseline Intravenous fluids (IV) | Order of IV fluids (NS, ½NS, D5NS, D5½NS, LR, D5WLR, Albumin 5%, 20%) in the 12 hours before check-in to the ward to 12 hours after check-in to the ward |
| Pre-alert IVF | Order of IV fluids (NS, ½NS, D5NS, D5½NS, LR, D5WLR, Albumin 5%, 20%) in the 12 hours before the alert |
| Antibiotics (number, %) |  |
| Baseline Antibiotics | antibiotics ordered in the 12 hours before check-in to the ward to 12 hours after check-in to the ward |
| Pre-alert Antibiotics | antibiotics ordered in the 12 hours before alert |

**Table S9:** Process Measures and post-alert physiologic variables.

| **Variable** | **Description** |
| --- | --- |
| Post-alert lactate | Reported in the 12 hours after the alert (number, %) |
|  | Highest value reported in the 12 hours after the alert |
| Post- alert blood culture (number, %) | Blood culture ordered in the 12 hours after alert |
| Post-alert respiratory culture (number, %) | Respiratory culture ordered in the 12 hours after alert |
| Post-alert urine culture (number, %) | Urine culture ordered in the 12 hours after alert |
| Post-alert body fluid culture (number, %) | Body fluid culture ordered in the 12 hours after alert |
| Antibiotics (number, %) | New antibiotics administered in the 12 hours after the alert |
|  | New antibiotics administered in the 3 hours after the alert |
| Post-alert systolic blood pressure | The lowest value in the 12 hours after the alert |
| Post-alert diastolic blood pressure | The lowest value in the 12 hours after the alert |
| Post-alert heart rate | The highest value in the 12 hours after the alert |
| Post-alert respiratory rate (RR) | Highest value RR in the 12 hours after the alert |

**Table S10:** Outcomes.

| **Variable** | | **Description** |
| --- | --- | --- |
| In-hospital mortality (number, %) | | Percentage of patients who die in the hospital up to 90 days |
| Hospital length of stay | | Stay in the hospital in days censored at 90 days |
| ICU admission | | Admission to ICU anytime during this hospital admission from check-in* to the ward up to 90 days; date and time, number of days in ICU |
| Post-alert ICU admission | | Transfer to ICU within 14 days of the alert  (number, %) |
| ICU-free days | | Number of days during the measurement period (maximum of 90 days) that the patient is both alive and free of mechanical ventilation |
| Incident renal replacement therapy (number, %) | | Renal replacement therapy (hemodialysis or continuous renal replacement therapy) any time this hospital admission after check-in to the ward up to 90 days |
| Post-alert renal replacement therapy (RRT) (number, %) | | Renal replacement therapy within 14 days from the alert |
| Vasopressor therapy (number, %) | | Vasopressors (norepinephrine, dopamine, epinephrine, dobutamine, phenylephrine, vasopressin) within 90 days of check-in; date and time of initiation |
| Post-alert vasopressors (number, %) | | Vasopressors within 14 days of the alert; date and time of initiation |
| Mechanical ventilation (number, %) | | Mechanical ventilation (except in the operating room) anytime this hospital admission after check-in to the ward to 90 days |
| Post-alert Mechanical ventilation (number, %) | | Mechanical ventilation (except in the operating room) within 14 days of the alert |
| Critical Care Rapid Response Team (CCRT) activation (number, %) | | CCRT activation during this hospital admission after check-in to the ward; date and time |
| Post-alert CCRT activation (number, %) | | CCRT activation within 14 days of the alert; date and time |
| Code Blue (number, %) | | Code Blue activation during this hospital admission after check-in to the ward  (number, %) Date and time |
| Post-alert Code Blue (number, %) | | Code Blue activation within 14 days of the alert; date and time |
| Post-alert antibiotics (number, %)  Antibiotic free-days up to 90 days | | Antibiotic ordered within 14 days of the alert  Number of days during the measurement period (maximum of 90 days) that the patient is both alive and free of antibiotics |
| Multidrug-resistant organism (number, %) | | A positive culture for multidrug-resistant Acinetobacter, Pseudomonas, bacteria with extended-spectrum beta-lactamases, methicillin-resistant Staphylococcus aureus, vancomycin-resistant enterococcus, carbapenem-resistant Klebsiella pneumoniae during 90 days from check-in |
| Post-alert MDROs (number, %) | | Starting from alert, positive culture for multidrug-resistant Acinetobacter, Pseudomonas, bacteria with extended-spectrum beta-lactamases, methicillin-resistant Staphylococcus aureus, vancomycin-resistant enterococcus, carbapenem-resistant Klebsiella pneumoniae up to 90 days |
| Clostridium difficile infection (number, %) | | Positive Clostridium difficile toxins by serology or polymerase chain reaction during 90 days from check-in |
| Alert Clostridium difficile infection (number, %) | | Start from alert: Positive Clostridium difficile toxins by serology or polymerase chain reaction during 90 days |
| **Electronic SOFA** | | Assessment of organ dysfunction using electronic data |
| Creatinine criterion | | The highest value of serum creatinine divided by the lowest creatinine from check-in to up to 90 days. If ≥ 2 then organ dysfunction is present; if < 2 then absent. |
| Bilirubin criterion | | The highest value of serum bilirubin divided by the lowest bilirubin from check-in to up to 90 days. If ≥ 2 then organ dysfunction is present; if < 2 then absent. |
| Lactate criterion | | The highest value of serum lactate divided by the lowest lactate from check-in to up to 90 days. If ≥ 2 then organ dysfunction is present; if < 2 then absent. |
| Platelet criterion | | The lowest platelet count divided by the highest platelet count from check-in to up to 90 days. If lowest platelet count is < 100 and ratio ≥ 2 then organ dysfunction is present; if count is > 100 or ratio < 2 then absent. |

*Check in to ward: ward admission; ward check out: ward discharg

**Table S11:** Justification for Sample Size calculation.

**Table S11 A:** A retrospective historical electronic data from 01July 2018 to 30 June 2019 was used to consider inputs for sample size calculation. The summary of the results are as follow.

| **Table of Alert by deaths** | | | |
| --- | --- | --- | --- |
|  | **Death** | | |
| **Alert (qSofa Alert)** | **Y** | **N** | **Total** |
| **Y** | **565** (8.16) | **6356** (91.84) | **6921 (**18.28) |
| **N** | **620** (2.00) | **30311** (98.00) | **30931 (**81.72) |
| **Total** | **1185** (3.13) | **36667** (96.87) | **37852 (**100.00) |

**Table S11 B:** Assumptions used for the sample size calculation.

|  | **Inputs** | **Justification** |
| --- | --- | --- |
| A) The implementation of qSOFA sepsis alert affects only patients with qSOFA sepsis alert | Yes | i.e. 6921 patients |
| B) Only half of patients with qSOFA sepsis alert have sepsis | Yes | 6921/2= 3461 |
| C) 90% of deaths in the sepsis alert group are sepsis related | Yes | Sepsis-related mortality=565*0.9=509  Non-sepsis related mortality=56 |
| D) Early intervention resulting from qSOFA sepsis alert will reduce the sepsis-related mortality 50% | 8.16% to 4.08% | Sepsis related mortality 509 deaths will be reduced by 50% to 255. |
| - Expected mortality rate before intervention is: | 3.13% | A total of 1185 deaths out of 37852 patients. |
| - Mortality rate expected to be reduced after intervention : | 2.46% | 620+255+56 deaths= 931  Then mortality rate =931/37852=2.46 |
| - Difference in mortality rate | 0.67% | Reduction in mortality after intervention |
| D) Expected intra-cluster correlation | 0.22 |  |
| Study Power | 90% |  |
| Level of significance | 5% |  |
| Sample Size | 62550 | An average of 1390 subjects per cluster with an average of 139 subjects per cluster per time period. |

qsofa: quick Sequential Organ Failure Assessment
